# Trade union strength and workers’ health and wellbeing across low-, middle-, and high-income countries: a repeated cross-sectional study of 69 countries

**DOI:** 10.64898/2026.11.27.26369126

**Authors:** Felix Forster, Jacques Wels

**Affiliations:** University College London (UCL), Unit for Lifelong Health and Ageing (LHA), London, UK; Université libre de Bruxelles (ULB), Health and Society Research Unit, Belgium

## Abstract

**Background:** Most research on the relationship between trade unions and workers’ health has focused on high-income countries. We address this gap by examining how trade union density and collective bargaining coverage are associated with workers’ self-rated health and life satisfaction across different economic settings, and how these associations have changed over time.

**Methods:** We analysed World Values Survey data on employees and self-employed workers, linked to OECD and ILO measures of national union density and collective bargaining coverage. We fitted ordinal regression models adjusted for individual and country-level characteristics, including interactions with survey year. We repeated analyses in low- and lower-middle-income countries (LMIC) and predicted outcomes at different levels of both indicators.

**Results:** Analyses included 114,586 individuals from 67 countries for self-rated health and 117,737 from 69 countries for life satisfaction. Higher union density was associated with better self-rated health and life satisfaction, with associations strengthening over time. Higher collective bargaining coverage was associated with worse outcomes, although associations weakened over time. Associations were stronger but less precise in LMIC. Model-based predictions for low-income countries showed better outcomes with higher union density in later years, but worse outcomes with greater bargaining coverage in earlier years.

**Discussion:** Higher union density was strongly associated with better health and wellbeing, particularly in low- and lower-middle-income countries, with associations strengthening over time. Despite limitations in data quality, these findings highlight the potential importance of trade unions strength as structural determinants of population health.

## Background

Recent evidence has shown that democracy at the workplace – i.e., collective negotiation and trade union membership – may have a positive impact on population health and wellbeing. ^1–4^ Epidemiological studies on this relationship have generally followed two methodological frameworks. The first comprises individual-level longitudinal studies that estimate the causal effects of trade union membership and company union presence on workers’ wellbeing outcomes. ^5,6^ The second uses cross-national comparative data to examine country-level differences in union density and coverage, typically focusing on European or OECD countries and considering both employed and non-employed populations. ^7,8^ A limitation of both approaches is their almost exclusive focus on high-income (HI) countries. ^9^ The present study addresses this gap by adopting the cross-sectional comparative approach – but extending the analysis to include low- and lower-middle-income countries (LMIC). In doing so, we aim to provide a broader understanding of the relationship between trade unions strength and workers’ wellbeing across a wider range of national contexts.

Trade union strength can be quantified by different measures. Trade union density (TUD), defined as the percentage of employees who are members of a trade union or representative organization, is the most commonly used measure. ^10–12^ In OECD countries, a steady decline in trade union membership was observed over the past forty years ^13^: union density mean was 30 percent in 1985, 20 percent in 2005 and around 15 percent in 2024. ^13^ However, moving beyond membership as a measure of union strength is important, as membership depends on individuals’ propensity to join a union ^14^ and institutional arrangements at the company or sectoral level. ^15,16^ Collective bargaining coverage (CBC), defined as the proportion of eligible employees covered by agreements negotiated at company, sectoral, cross-sectoral, or national level, provides a complementary indicator. ^17,18^ Although bargaining coverage has also declined across OECD countries, from approximately 47% in 1985 to 34% in 2023–24, its evolution differs from that of union density. ^13^ Important differences also exist across countries, with relatively stable unionization in Belgium but substantial declines in Finland, as well as discrepancies between administrative and survey estimates in some settings. ^15^ These differences suggest that union membership and collective bargaining coverage capture distinct dimensions of labour representation.

These measures are often used separately. Using three waves of the European Social Survey, Reynolds and Buffel found that higher country-level union density was associated with fewer depressive symptoms. ^7^ Moving beyond union density, Sochas and Reeves, using the European Values Survey across 33 OECD countries, showed that stronger and more inclusive collective bargaining institutions were associated with reduced health inequalities between employed and unemployed individuals, primarily through better health among the unemployed. ^8^ However, cross-national evidence from non-OECD countries remains sparse, and few studies have systematically addressed low- and lower-middle-income settings. Using World Bank enterprise data, Bryson and Tanaka found that trade unions were present in approximately 30% of firms in developing countries, with a mean firm-level union density of 18%. ^19^ Yet substantial differences exist across countries, not only in unionization rates but also in union functions, institutional arrangements, and freedom of association.

A global approach to collective negotiation requires to question the indicators that are traditionally used in high income countries. Union density, for instance, is traditionally calculated as the ratio of unionized workers over the employed population. Such a definition implies that only employed workers have access to unions and that the self-employed population would not be covered by any form of collective bargaining. Whilst this is true in many high-income countries, where initiatives by main trade unions ^20^ or specific forms of unions (e.g., community unions) ^16,21^ to support the self-employed – and also the less traditionally unionized workers – exist but are not widespread, a large part of the workforce in LMIC is self-employed ^22,23^ and *de facto* out of scope of the traditional union density indicator.

More fundamentally, this questions the very nature of trade unions. Rather than being understood strictly as institutions embedded in an employment relationship, unions can also be conceptualized as associational forms of collective action among workers operating outside standard employment relations. Research on informal employment ^24^ has documented how street vendors, waste pickers, transport workers, and home-based workers develop collective organizations that perform functions traditionally associated with trade unions ^25,26^. These organizations negotiate over regulation, access to space and infrastructure, pricing conditions, and social protection, exercising forms of collective bargaining without an employer in the classical sense. Conventional indicators of union strength may therefore fail to capture important dimensions of workers’ collective representation in these settings.

Using data from the World Values Survey (WVS) – one of the largest cross-national surveys of values and social attitudes – this study aims to investigate how different dimensions of collective labour representation, beyond individual trade union membership, are associated with workers’ self-rated health and life satisfaction across up to 69 countries, and how these associations have changed over time. Although previous studies using WVS data have examined trade union membership, union density, and life satisfaction ^27–29^, to our knowledge, none has investigated both trade union density and collective bargaining coverage in relation to workers’ health and wellbeing over time, particularly in LMIC.

## Data and Methods

### Study population

We analysed publicly available data from the World Values Survey, which contains data collected in seven waves between 1981 and 2022 from a large number of countries. ^30^ Not all surveys, i.e., the separate waves conducted in different countries, used completely identical questionnaires. We excluded surveys that did not ask for individual union membership, occupation, employment status, gender, or the corresponding outcome. In addition, to avoid bias related to the COVID-19 pandemic, we excluded data collected after 2019. We restricted analyses to the working population, i.e., full-time and part-time employees, as well as self-employed (see flowchart in Figure S1 in the supplement). In addition to individual-level WVS data, we included country-specific information, as described below.

### Exposure

Our exposures of interest were trade union density and collective bargaining coverage. Both TUD and CBC were country-level variables that varied by year.

We used data from the Institutional Characteristics of Trade Unions, Wage Setting, State Intervention and Social Pacts (ICTWSS) database maintained by the Organisation for Economic Co-operation and Development (OECD) and the Amsterdam Institute for Advanced Labour Studies and the Hugo Sinzheimer Institute (AIAS-HSI) ^31^, as well as data from the Industrial Relations Data (IRdata) database by the International Labour Organization (ILO). ^32^

TUD was defined as the proportion of employees who are members of a trade union, excluding self-employed, retired, students, and unemployed, whenever possible. ^31,32^ CBC was defined as the proportion of employees covered by collective bargaining, excluding workers who are legally not allowed to collectively bargain, whenever possible. ^31,32^

If values were missing for the years of the WVS surveys, we used the closest available value. If two values were equally close (same number of years into the past and the future), we used the most recent, based on the assumption that estimation tends to improve. Countries without TUD or CBC estimates were excluded. If both ICTWSS and ILO estimates were available for the same year, we used ICTWSS values.

### Outcome

Our primary outcome was self-rated health (“All in all, how would you describe your state of health these days?”). ^30^ Some WVS waves used five answer options (Very good, Good, Fair, Poor, Very poor) and some used only four, leaving out “Very poor”. We therefore used the four-category variant and combined “Poor” and “Very poor”, when necessary.

Our secondary outcome was life satisfaction (“All things considered, how satisfied are you with your life as a whole these days?”). ^30^ The item was measured with a 10-point Likert scale from “Dissatisfied” to “Satisfied”, which we compressed to five categories by systematically combining two options, in order to allow analysis with ordinal regression.

### Covariates

We additionally considered individual-level covariates from WVS data: reported gender (male, female), age group (<20 years, 20-29, 30-39, 40-49, 50-59, 60+), trade union membership (yes, no), and occupation (primarily non-manual, primarily manual, agricultural). Variable “occupation” harmonised two different sets of answer categories from different surveys. As part of the harmonisation process, we assigned “primarily non-manual” to (among others) sales workers and managers of small establishments, and “primarily manual” to (among others) service workers.

Country-level data from The World Bank was used to define country income (low, lower-middle, upper-middle, and high income) ^33^ and income inequality (Gini index). ^34^ For the Gini index, we used the closest available value within countries when values were missing, similar to TUD/CBC. Like all country-level variables, income classifications are available on a yearly resolution and change over time.

Data from the Varieties of Democracy (V-Dem) Project was used to describe the state of democracy by country. ^35,36^ We used the liberal democracy index, which measured the liberal components of democracy (equality before the law and individual liberty, judicial and legislative constraints on the executive) in addition to the basic electoral components of democracy.

### Statistical analysis

We used two successive methods. First, we applied ordinal regression (cumulative link models with logit link) with separate cut-points for each country ^37^, assuming that a similar value of the underlying continuous variable might be classified differently by country. Models were adjusted in four steps: 1) by the exposure of interest (TUD or CBC) and its interaction with survey year, 2) additionally by gender, age group, and occupation, 3) additionally by individual trade union membership and the second exposure of interest (TUD/CBC), and 4) additionally by democracy and Gini indices. The fully adjusted model was refit using only data from low- and lower-middle-income countries (classification for year of data collection). The models were fit for all four combinations of exposure (TUD, CBC) and outcome (self-rated health, life satisfaction).

Continuous variables were transformed to increase interpretability of estimates. TUD, CBC, democracy index, and Gini index were standardised. Survey year was transformed to the number of decades since the start of the study. Parameter estimates of the exposure of interest and its interaction with survey year were combined by fixing survey year to values 1985, 1995, 2005, and 2015 and calculating estimate and standard error ^38^ for the exposure variable.

Second, to evaluate the implications of the parameter estimates for LMIC, we predicted outcome values based on the actual data, while changing the values of the exposure of interest (TUD or CBC, but not both). First, we created input data for prediction from the observed data by changing the observed value of TUD or CBC to a fixed value. We used 20%, 40%, 60%, and 80%, creating four separate sets of predictions. Next, we used these datasets to calculate predictions, which were vectors of numbers, equal in length to the number of outcome categories, for each participant describing their probability of being in a certain outcome category. Next, we sampled 100 times from these probabilities, creating 100 different realisations of the predicted probabilities, i.e., 100 variables similar to the observed outcome. Next, in order to enable visualisation, we calculated the mean of each variable within country and wave by replacing each category with an integer from 1 to n (1 to 4 for self-rated health, 1 to 5 for life satisfaction) with higher integers referring to worse health/satisfaction, i.e., an expected outcome value. We applied the same summary to the observed outcome for comparison. Finally, to describe sampling variance, we summarised the 100 predicted expected outcome values by mean, as well as 2.5% and 97.5% quantiles. Predictions were created for the fully adjusted model as well as the fully adjusted model restricted to LMIC.

All analyses were conducted in R version 4.6.1. ^39^ Ordinal regression models were conducted with the “ordinal” package. ^37^ The analysis code is available on GitHub. ^40^

### Sensitivity analyses

In addition, we conducted sensitivity analyses where we did not exclude surveys after 2019, where we restricted to surveys for which both TUD and CBC were available for the year of data collection, where we included employees only, and where we weighted each country equally. Because two-way interactions may bias the estimates when the covariates are linked to the outcome ^41^ , we additionally stratified the analyses by wave and compared the trend to the main models.

## Results

### Study population

In total, 114,586 and 117,737 individuals participated in the included surveys for self-rated health and life satisfaction, respectively (see flowchart in Figure S1). Due to individual missing values, 350-6,653 (self-rated health) and 891-8,060 (life satisfaction) participants were excluded across the different models. The datasets for self-rated health and life satisfaction included data from 150 and 153 surveys, i.e., separate waves in different countries, conducted in 67 and 69 countries, respectively, with life satisfaction data additionally including the Dominican Republic and Singapore. For both self-rated health and life satisfaction, 14 surveys from 10 countries were conducted in countries classified as having low (but not lower-middle) income at the time.

Lowest outcome categories, i.e., poor or very poor self-rated health and very low life satisfaction, were relatively rare with proportions of 4.6% and 5.0%, respectively (Table 1). Overall, the distribution of gender and age reasonably reflected a working population, and about 10% of individuals worked in agriculture, while about 20% were self-employed. The proportion of trade union membership across all individuals was 22%. Summary statistics of country-level variables depended on both the actual values as well as the number of participants per country and were used for variable standardisation. Summaries for datasets used for models of self-rated health as well as models for life satisfaction are almost identical as most individuals are part of both datasets. See Table S1 in the supplement for country-specific descriptives.

**Table 1:** Descriptive statistics by individual participants, separate for data considered for models of self-rated health and life satisfaction. Trade union density, collective bargaining coverage, democracy index, and Gini index are country-level variables. Gini index is missing completely for New Zealand and Singapore.

| Variable | Self-rated health |  | Life satisfaction |  |
|---|---|---|---|---|
|  | Missing | N = 114,586 | Missing | N = 117,737 |
|  | n (%) | n (%) | n (%) | n (%) |
| Self-rated health |  |  |  |  |
| Very good | 350 (0.3%) | 27,780 (24.3%) | - | - |
| Good |  | 51,632 (45.2%) |  | - |
| Fair |  | 29,561 (25.9%) |  | - |
| Poor or very poor |  | 5,263 (4.6%) |  | - |
| Life satisfaction |  |  |  |  |
| Very high | - | - | 891 (0.8%) | 28,197 (24.1%) |
| High |  | - |  | 43,268 (37.0%) |
| Medium |  | - |  | 28,191 (24.1%) |
| Low |  | - |  | 11,399 (9.8%) |
| Very low |  | - |  | 5,791 (5.0%) |
| Female | 64 (0.1%) | 48,589 (42.4%) | 64 (0.1%) | 49,874 (42.4%) |
| Age group |  |  |  |  |
| <20 | 111 (0.1%) | 2,341 (2.0%) | 118 (0.1%) | 2,520 (2.1%) |
| 20-29 |  | 25,777 (22.5%) |  | 26,699 (22.7%) |
| 30-39 |  | 32,205 (28.1%) |  | 33,090 (28.1%) |
| 40-49 |  | 28,601 (25.0%) |  | 29,266 (24.9%) |
| 50-59 |  | 18,415 (16.1%) |  | 18,791 (16.0%) |
| 60+ |  | 7,136 (6.2%) |  | 7,253 (6.2%) |
| Occupation |  |  |  |  |
| Primarily non-manual | 4,400 (3.8%) | 54,391 (49.4%) | 4,550 (3.9%) | 55,916 (49.4%) |
| Primarily manual |  | 44,363 (40.3%) |  | 45,735 (40.4%) |
| Agricultural |  | 11,432 (10.4%) |  | 11,536 (10.2%) |
| Employment status |  |  |  |  |
| Full-time employed | 0 (0.0%) | 76,429 (66.7%) | 0 (0.0%) | 78,712 (66.9%) |
| Part-time employed |  | 15,351 (13.4%) |  | 15,928 (13.5%) |
| Self-employed |  | 22,806 (19.9%) |  | 23,097 (19.6%) |
| Trade union membership | 734 (0.6%) | 25,188 (22.1%) | 728 (0.6%) | 25,849 (22.1%) |
|  | n (%) | Mean (SD) | n (%) | Mean (SD) |
| Trade union density | 0 (0.0%) | 25 (16.0) | 0 (0.0%) | 25 (16.6) |
| Collective bargaining coverage | 0 (0.0%) | 35 (27.0) | 0 (0.0%) | 34 (27.3) |
| Democracy index | 0 (0.0%) | 52 (26.7) | 0 (0.0%) | 51 (26.6) |
| Gini index | 1,310 (1.1%) | 39 (9.2) | 2,174 (1.8%) | 39 (9.4) |

Figure 1. shows TUD and CBC values for the 154 surveys included in the study. Up to values of about 30%, many points were close to the dotted line where TUD and CBC have the same value. In total, more points fell on the right side of the dotted line, where CBC is higher than TUD. However, CBC values substantially larger than their corresponding TUD appeared only above a TUD of approximately 15%. Most TUD and CBC values above 50% were from upper-middle- or high-income countries. For many points, either TUD or CBC was not available for the year of data collection. Figure S2 in the supplement shows that less data was available for CBC as well as for LMIC.

**Figure 1:**
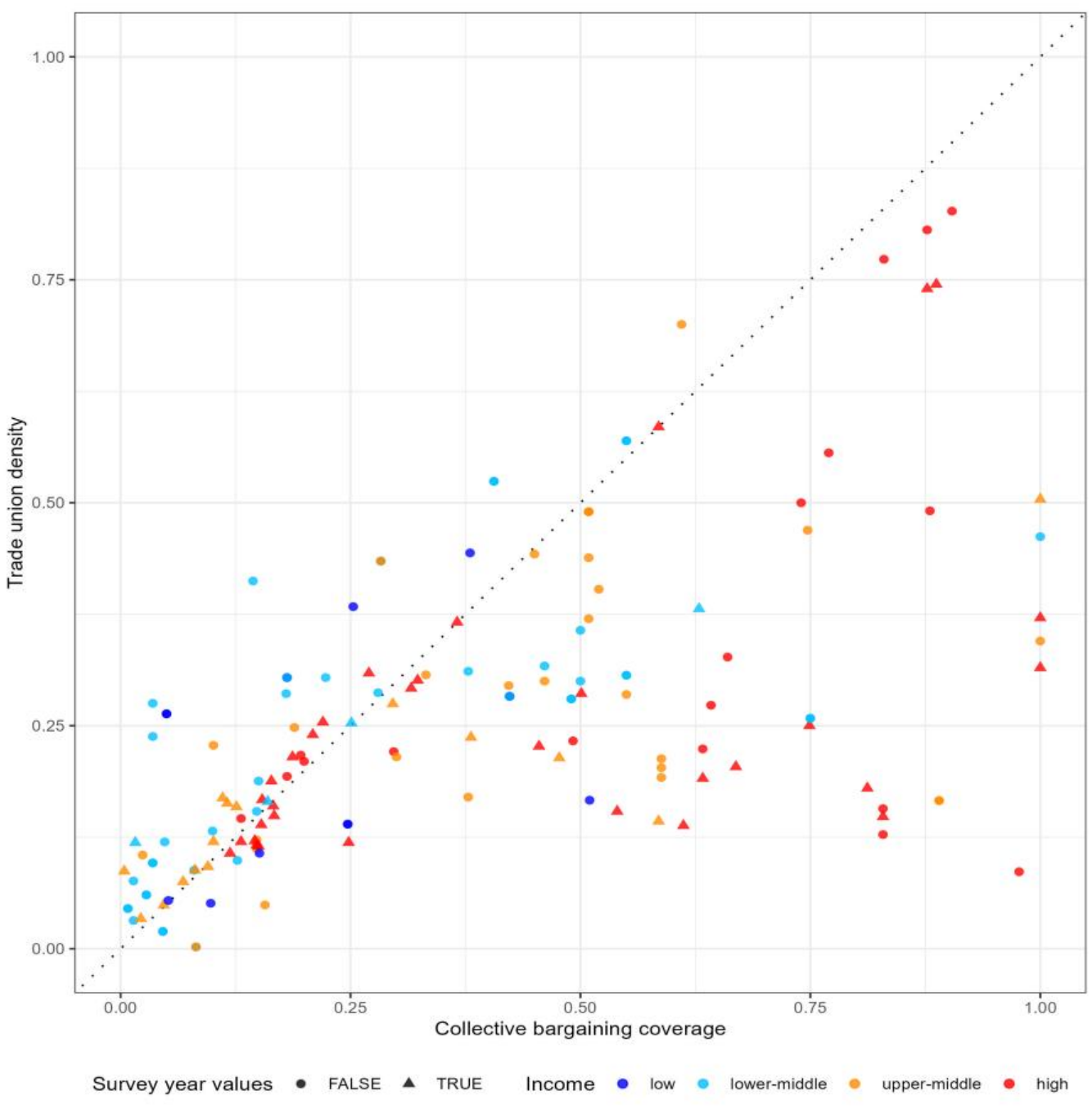
Combinations of trade union density (TUD) and collective bargaining coverage (CBC) for every survey included in the study. Colour indicates country income. Triangles indicate that both TUD and CBC were available for the year of data collection, while dots indicate that at least one of TUD/CBC was not available for the year of data collection and was instead taken from another year.

### Estimates

Figure 2. shows the proportional odds ratios (ORs) for the associations of TUD and CBC with self-rated health and life satisfaction. These ORs represent the change in the odds of reporting a given outcome category or worse, rather than any better category. Effect estimates are expressed as ORs per one-standard-deviation increase in TUD or CBC, corresponding to approximately 16 and 27 percentage points, respectively (see descriptives in Table 1).

**Figure 2:**
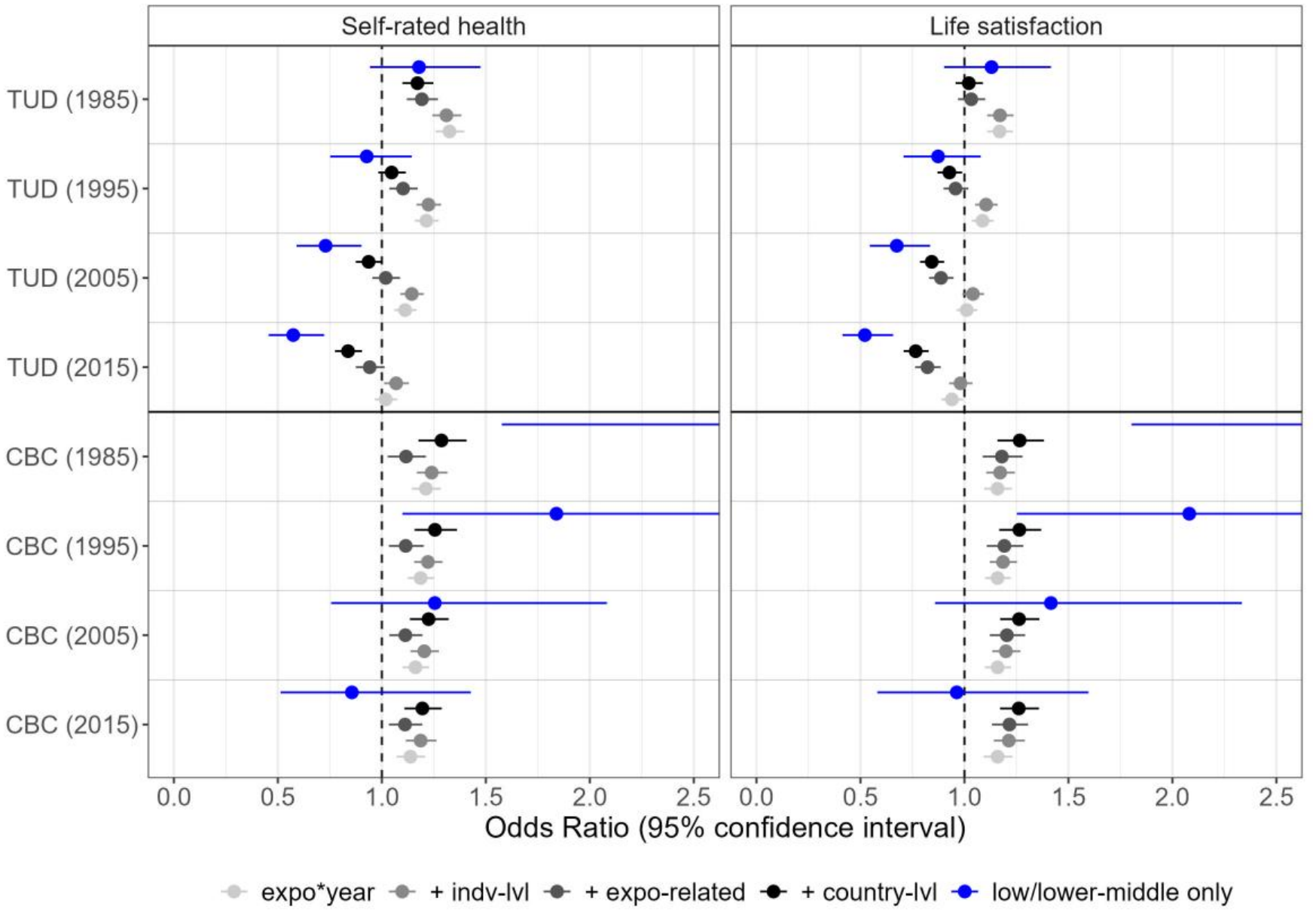
Effect estimates for trade union density (TUD) and collective bargaining coverage (CBC) from ordinal regression models. Due to interactions with survey year, estimates and standard errors have been calculated for the years 1985, 1995, 2005, and 2015. Estimates coloured in grey/black include all countries and darker colour indicates adjustment for more factors (“expo*year”: only includes main exposure, TUD or CBC, and its interaction with survey year; “+ indv-lvl”: additionally includes individual-level covariates gender, age group, and occupation; “+ expo-related”: additionally includes covariates related to trade unions, namely individual trade union membership and the other exposure; “+ country-lvl”: additionally includes country-level covariates democracy index and Gini index). Blue estimates are from fully adjusted models that only include data from low- and lower-middle-income countries. The estimated values are available in Table S2.

Effect estimates for TUD on self-rated health started above 1 indicating worse health ratings associated with increased TUD, but estimates moved towards “better health ratings” with additional adjustment and with time. In contrast, increasing CBC was associated with worse self-rated health in our models. Models for life satisfaction generally showed the same patterns, while models that only use data from LMIC showed stronger associations in the same directions. Additionally, interval estimates were considerably larger in LMIC models, especially for CBC.

### Predictions

To better understand the implications of the estimates shown above for low-income countries, we predicted outcome distributions for study participants across different hypothetical exposure scenarios. **Figure 3** shows predictions of expected self-rated health (columns 1 & 3) and life satisfaction (columns 2 & 4) in low-income countries for varying values of TUD (columns 1 & 2) and CBC (columns 3 & 4).

**Figure 3:**
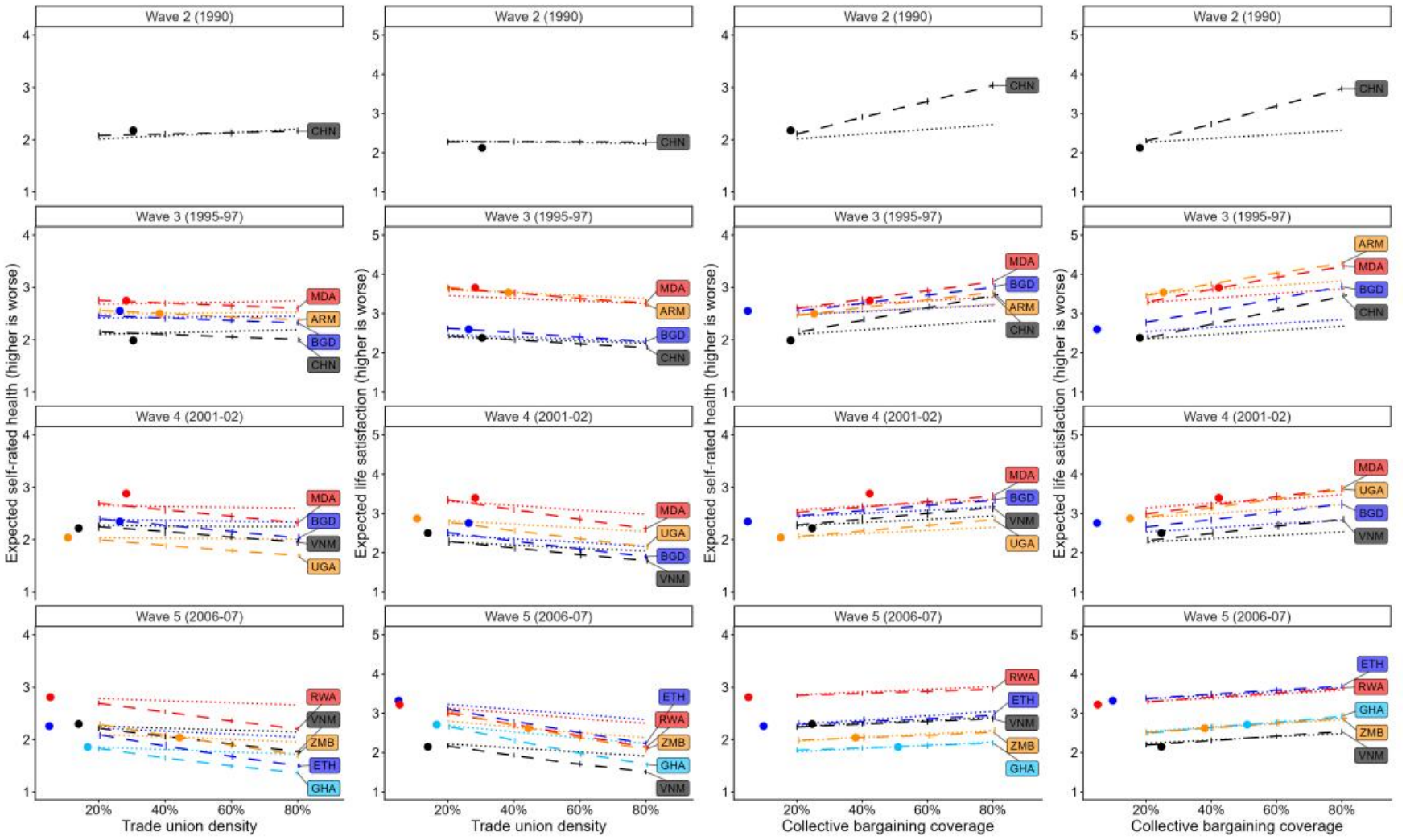
Predicted (lines) and observed (dots) expected self-rated health (columns 1 & 3) and life satisfaction (columns 2 & 4) for different values of trade union density (column 1 & 2) and collective bargaining coverage (columns 3 & 4) by country (low-income only) and study wave in fully adjusted models including all countries (dotted lines) and including low- and lower-middle-income countries only (dashed lines). Line ranges show 2.5 and 97.5% quantiles for predicted expected outcomes, for exposure values of 20%, 40%, 60%, and 80%. Lines only connect these four sets of predictions. ARM: Armenia, BGD: Bangladesh, CHN: China, ETH: Ethiopia, GHA: Ghana, MDA: Moldova, RWA: Rwanda, UGA: Uganda, VNM: Vietnam, ZMB: Zambia

The observed values (dots) aligned well with the predicted values (lines) for most surveys. While predictions in earlier years showed no real trend for varying values of TUD, increasing TUD in later years led to predictions of better self-rated health and life satisfaction. In contrast, in earlier years, increasing CBC was related to predictions of worse health/satisfaction, while the strength of the relation was predicted to get weaker over time. When comparing predictions with models that includes all countries (dotted lines), models with LMIC data only (dashed lines) show stronger trends for both TUD and CBC. For China (CHN) in wave 2, e.g., predictions of self-rated health for a TUD of 20% and 80% were 2.01 and 2.21, respectively, based on models using data from all countries, and 2.08 and 2.17 based on LMIC models, while for Rwanda (RWA) in wave 5, predictions were 2.79 and 2.66 in models based on all countries, and 2.69 and 2.20 in LMIC models. In contrast, for CBC, predictions of self-rated health for China in wave 2 for a CBC of 20% and 80% were 2.02 and 2.29 in models based on all countries, and 2.11 and 3.04 in LMIC models, while for Rwanda in wave 5, predictions were 2.85 and 3.01 for models based on all countries, and 2.84 and 2.96 in LMIC models. Predictions for lower-middle-income countries showed similar trends (see Figure S3).

### Sensitivity analysis

Including post-2019 surveys (Figure S4) generally produced estimates similar to the main analyses, although associations between TUD and self-rated health were less favourable, particularly in LMIC. Estimates for CBC were more variable in LMIC, with wider confidence intervals and associations differing between self-rated health and life satisfaction.

Restricting analyses to surveys with contemporaneous TUD and CBC measurements (Figure S5) also produced less favourable estimates for TUD, with no protective associations observed. For CBC, associations with life satisfaction remained broadly similar, whereas those with self-rated health became protective. However, this restriction substantially altered the sample composition, disproportionately excluding low- and lower-middle-income countries, where the main analyses showed stronger positive associations between TUD and health and wellbeing.

Restricting to employees only and including country weights lead to similar estimates than the main models (see Figures S6 & S7). The trend in models stratified by wave was relatively similar to the trend estimated by the interaction term (see Figures S8a-d).

## Discussion

We investigated the relationship of trade union density and collective bargaining coverage with self-rated health and life satisfaction in cross-sectional data from 69 countries, including low- and lower-middle-income countries. We found that higher trade union density is associated with better outcome ratings and these became stronger over time. In contrast, collective bargaining coverage is associated with worse outcome ratings. In LMIC, associations were estimated to be stronger. In predictions for low-income countries, the association for CBC became weaker over time. Similar results were observed for self-rated health and life satisfaction.

The differential relationship observed between union density and collective bargaining coverage can be explained by three factors. First, unions may influence wellbeing beyond collective agreement ^42,43^. While collective agreements primarily focus on economic benefits, unions also fulfil broader social and political functions, including fostering solidarity, providing collective voice, and advocating for policies that promote wellbeing. Second, high bargaining coverage does not necessarily indicate a robust bargaining system, and coverage can remain high even where union bargaining power is limited ^44,45^. Third, the quality of bargaining institution matters more than coverage alone. Two countries with similar bargaining coverage rates may have very different systems in terms of union independence, bargaining coordination, enforcement, employer cooperation and the quality of negotiated agreements. For instance, it was found that the relationship between union membership and job satisfaction varies according to national industrial relations institutions ^46^. By contrast, union density may better reflect the organisational strength of unions and their capacity to represent workers’ interests. This distinction may be particularly important in low- and middle-income countries, where widespread informal employment, limited social protection, and weak enforcement of labour regulations may reduce the effectiveness of formal collective bargaining agreements, making unions’ broader social and political roles especially relevant to health and wellbeing.

Despite being the first study to assess such relationships, it has several limitations. First, the study uses cross-sectional data, which limits causal inference. Although using population-level rather than individual-level data reduces concerns about selection bias associated with union membership, the study design does not allow us to establish whether the observed relationships are causal. Second, TUD and CBC values were not available for the data collection years for a considerable number of countries and years, with data quality worse in LMIC. We imputed data based on other available values, primarily from later years. Sensitivity analyses restricting the analyses to surveys for which TUD and CBC values were available for the same year, changed the effect estimates but dropped LMIC. Third, our sensitivity analyses did not show difference when focusing on the full working population (including the self-employed) or the employed population. However, TUD and CBC data are mainly focusing on employees, and this might underestimate the strength of unions, particularly in countries with a high self-employed workforce.

## Conclusion

Higher trade union density is strongly associated with better self-rated health and life satisfaction, particularly in low- and lower-middle-income countries, with associations strengthening over time. By contrast, collective bargaining coverage was associated with poorer outcomes, suggesting that these indicators capture distinct dimensions of workers’ representation. Our findings highlight the importance of trade unions for understanding population health and wellbeing, particularly in lower-income settings, and the need to reconsider how collective labour representation is measured across different economic and institutional contexts.

## Data Availability

The World Values Survey (WVS) data can be accessed and downloaded from the World Values Survey Association's website (https://www.worldvaluessurvey.org/). Access is free but registration and acceptance of the data-use conditions may be required for certain datasets.
The ICTWSS database (Institutional Characteristics of Trade Unions, Wage Setting, State Intervention and Social Pacts) is freely available through the OECD/AIAS data repository (https://www.oecd.org/en/data/datasets/oecdaias-ictwss-database.html).
The ILO Industrial Relations Data (IRdata) are freely available on the website of the International Labour Organization (https://ilostat.ilo.org/databases/irdata/description/).
Data by The World Bank are freely available on their website (income classification: https://datahelpdesk.worldbank.org/knowledgebase/articles/906519-world-bank-country-and-lending-groups, GINI index: https://data.worldbank.org/indicator/SI.POV.GINI)
The V-Dem data is freely available and can be downloaded from their website (https://doi.org/10.23696/vdemds26).
For further information on how to download the data used for this analysis, see also the analysis code available via https://doi.org/10.5281/zenodo.24526077.

## Declarations

### Author contributions

Conceptualization: JW, FF; Data Curation: FF; Formal Analysis: FF; Funding Acquisition: JW; Investigation: FF, JW; Methodology: FF, JW; Project Administration: FF, JW; Resources: FF, JW; Software: FF; Supervision: FF; Validation: JW; Visualization: FF; Writing – Original Draft Preparation: FF, JW; Writing – Review & Editing: JW, FF.

### Data availability

The World Values Survey (WVS) data can be accessed and downloaded from the World Values Survey Association’s website (https://www.worldvaluessurvey.org/). Access is free but registration and acceptance of the data-use conditions may be required for certain datasets.

The ICTWSS database (Institutional Characteristics of Trade Unions, Wage Setting, State Intervention and Social Pacts) is freely available through the OECD/AIAS data repository (https://www.oecd.org/en/data/datasets/oecdaias-ictwss-database.html).

The ILO Industrial Relations Data (IRdata) are freely available on the website of the International Labour Organization (https://ilostat.ilo.org/databases/irdata/description/).

Data by The World Bank are freely available on their website (income classification: https://datahelpdesk.worldbank.org/knowledgebase/articles/906519-world-bank-country-and-lending-groups, GINI index: https://data.worldbank.org/indicator/SI.POV.GINI)

The V-Dem data is freely available and can be downloaded from their website (https://doi.org/10.23696/vdemds26).

For further information on how to download the data used for this analysis, see also the analysis code available via https://doi.org/10.5281/zenodo.24526077.

### Declaration of generative AI use

The authors did not use generative AI for this study.

