## Supplement for "Trade union strength and workers’ health and wellbeing across low-, middle-, and high-income countries: a repeated cross-sectional study of 69 countries"

Felix Forster[1], Jacques Wels[1,2]

[1] University College London (UCL), Unit for Lifelong Health and Ageing (LHA), London, UK

[2] Université libre de Bruxelles (ULB), Health and Society Research Unit, Belgium

### Flowchart

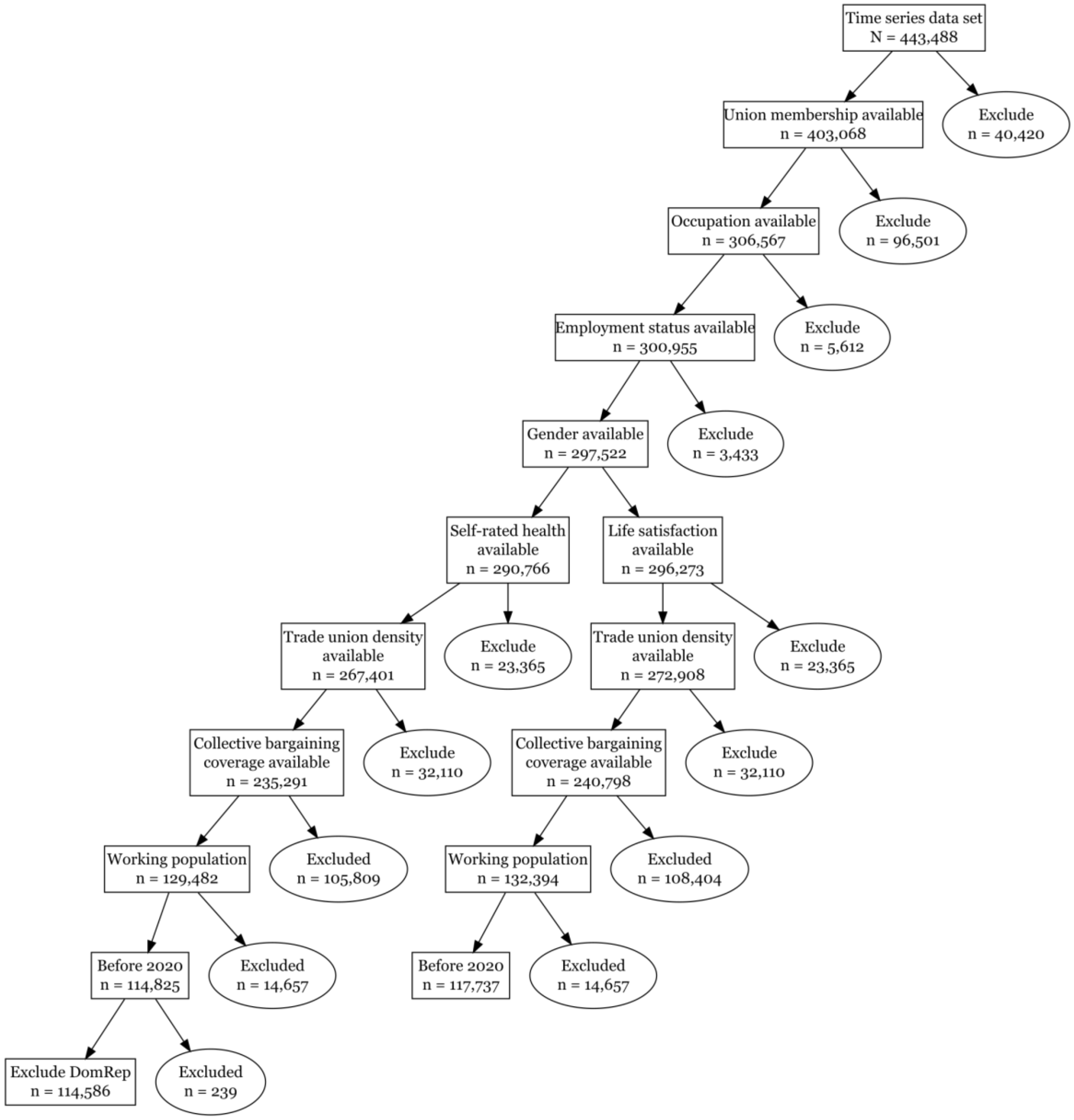


Figure S1: Flowchart of data processing. The branching describes the creation of the datasets used for models of outcome self-rated health (left) and life satisfaction (right)

### Country-specific descriptives

Table S1: Descriptives by country and wave. Income: low (L), lower-middle (LM), upper-middle (UM), or high (H) income country; n: number of included participants; SRH: self-rated health; LS: Life satisfaction; TUD: trade union density; CBC: collective bargaining coverage; TUM: proportion of trade union membership among included participants; %Self: proportion of self-employed among all included participants; %Self (TU): proportion of self-employed among included participants who are trade union members

| Wave (Year) | Income | n | In SRH data? | In LS data? | TUD | CBC | TUM | %Self | %Self (TU) |
|---|---|---|---|---|---|---|---|---|---|
| Albania | | | | | | | | | |
| 3 (1998) | LM | 475 | yes | yes | 52% | 41% | 15% | 52% | 22% |
| 4 (2002) | LM | 565 | yes | yes | 52% | 41% | 14% | 35% | 20% |
| Argentina | | | | | | | | | |
| 1 (1984) | UM | 585 | yes | yes | 49% | 51% | 15% | 22% | 13% |
| 2 (1991) | UM | 573 | yes | yes | 49% | 51% | 2% | 34% | 0% |
| 3 (1995) | UM | 515 | yes | yes | 49% | 51% | 9% | 30% | 7% |
| 4 (1999) | UM | 652 | yes | yes | 44% | 51% | 4% | 32% | 13% |
| 5 (2006) | UM | 562 | yes | yes | 37% | 51% | 11% | 22% | 17% |
| 7 (2017) | H | 607 | yes | yes | 27% | 64% | 10% | 37% | 29% |
| Armenia | | | | | | | | | |
| 3 (1997) | L | 948 | yes | yes | 38% | 25% | 100% | 21% | 22% |
| Australia | | | | | | | | | |
| 1 (1981) | H | 664 | yes | yes | 49% | 88% | 31% | 0% | 0% |
| 3 (1995) | H | 1162 | yes | yes | 33% | 66% | 33% | 5% | 2% |
| 5 (2005) | H | 862 | yes | yes | 22% | 63% | 28% | 17% | 6% |
| 7 (2018) | H | 985 | yes | yes | 14% | 61% | 26% | 13% | 3% |
| Bangladesh | | | | | | | | | |
| 3 (1996) | L | 604 | yes | yes | 26% | 5% | 9% | 36% | 25% |
| 4 (2002) | L | 457 | yes | yes | 26% | 5% | 24% | 39% | 41% |
| 7 (2018) | LM | 523 | yes | yes | 12% | 2% | 16% | 58% | 49% |
| Bosnia and Herzegovina | | | | | | | | | |
| 3 (1998) | LM | 594 | yes | yes | 30% | 50% | 50% | 10% | 7% |
| 4 (2001) | LM | 541 | yes | yes | 36% | 50% | 13% | 6% | 0% |
| Brazil | | | | | | | | | |
| 2 (1991) | UM | 1014 | yes | yes | 21% | 59% | 9% | 19% | 14% |
| 3 (1997) | UM | 652 | yes | yes | 19% | 59% | 25% | 13% | 4% |
| 5 (2006) | UM | 778 | yes | yes | 20% | 59% | 24% | 36% | 18% |
| 7 (2018) | UM | 845 | yes | yes | 14% | 58% | 17% | 33% | 15% |
| Bulgaria | | | | | | | | | |
| 3 (1997) | LM | 505 | yes | yes | 31% | 38% | 25% | 4% | 0% |
| 5 (2006) | UM | 441 | yes | yes | 17% | 38% | 13% | 3% | 0% |
| Canada | | | | | | | | | |
| 4 (2000) | H | 1077 | yes | yes | 30% | 32% | 21% | 8% | 3% |
| 5 (2006) | H | 1155 | yes | yes | 29% | 32% | 35% | 12% | 4% |
| Chile | | | | | | | | | |
| 2 (1990) | LM | 788 | yes | yes | 15% | 15% | 10% | 26% | 16% |
| 3 (1996) | UM | 514 | yes | yes | 12% | 15% | 23% | 23% | 11% |

| Wave (Year) | Income | n | In SRH data? | In LS data? | TUD | CBC | TUM | %Self | %Self (TU) |
|---|---|---|---|---|---|---|---|---|---|
| 4 (2000) | UM | 649 | yes | yes | 11% | 15% | 5% | 27% | 6% |
| 5 (2006) | UM | 491 | yes | yes | 12% | 15% | 21% | 18% | 17% |
| 7 (2018) | H | 691 | yes | yes | 16% | 17% | 23% | 8% | 10% |
| China | | | | | | | | | |
| 2 (1990) | L | 854 | yes | yes | 30% | 18% | 2% | 25% | 6% |
| 3 (1995) | L | 1242 | yes | yes | 30% | 18% | 21% | 16% | 4% |
| 4 (2001) | LM | 778 | yes | yes | 30% | 18% | 7% | 5% | 2% |
| 5 (2007) | LM | 1358 | yes | yes | 30% | 22% | 11% | 5% | 3% |
| 7 (2018) | UM | 1831 | yes | yes | 44% | 45% | 14% | 14% | 4% |
| Colombia | | | | | | | | | |
| 3 (1997) | LM | 1559 | NO | yes | 4% | 1% | 8% | 22% | 15% |
| 3 (1998) | LM | 1815 | yes | yes | 4% | 1% | 9% | 19% | 5% |
| 7 (2018) | UM | 808 | yes | yes | 5% | 16% | 27% | 30% | 27% |
| Croatia | | | | | | | | | |
| 3 (1996) | UM | 608 | yes | yes | 70% | 61% | 44% | 14% | 4% |
| Cyprus | | | | | | | | | |
| 5 (2006) | H | 606 | yes | yes | 58% | 58% | 24% | 19% | 8% |
| 7 (2019) | H | 581 | yes | yes | 37% | 37% | 23% | 18% | 18% |
| Czechia | | | | | | | | | |
| 3 (1998) | UM | 623 | yes | yes | 31% | 33% | 26% | 8% | 1% |
| Dominican Republic | | | | | | | | | |
| 3 (1996) | LM | 239 | NO | yes | 9% | 8% | 24% | 17% | 29% |
| Egypt | | | | | | | | | |
| 5 (2008) | LM | 1056 | yes | yes | 28% | 4% | 2% | 0% | 0% |
| 7 (2018) | LM | 597 | yes | yes | 24% | 4% | 4% | 19% | 4% |
| El Salvador | | | | | | | | | |
| 3 (1999) | LM | 612 | yes | yes | 12% | 5% | 3% | 35% | 20% |
| Estonia | | | | | | | | | |
| 3 (1996) | LM | 656 | yes | yes | 29% | 28% | 18% | 12% | 2% |
| Ethiopia | | | | | | | | | |
| 5 (2007) | L | 697 | yes | yes | 5% | 10% | 29% | 42% | 47% |
| Finland | | | | | | | | | |
| 3 (1996) | H | 475 | yes | yes | 77% | 83% | 69% | 8% | 5% |
| 5 (2005) | H | 515 | yes | yes | 74% | 88% | 69% | 8% | 3% |
| France | | | | | | | | | |
| 5 (2006) | H | 500 | yes | yes | 9% | 98% | 15% | 12% | 14% |
| Georgia | | | | | | | | | |
| 5 (2009) | LM | 508 | yes | yes | 41% | 14% | 9% | 41% | 2% |
| Germany | | | | | | | | | |
| 3 (1997) | H | 1081 | yes | yes | 25% | 75% | 25% | 4% | 1% |
| 5 (2006) | H | 881 | yes | yes | 19% | 63% | 18% | 10% | 2% |
| 7 (2018) | H | 871 | yes | yes | 15% | 54% | 17% | 7% | 4% |
| Ghana | | | | | | | | | |
| 5 (2007) | L | 893 | yes | yes | 17% | 51% | 27% | 73% | 61% |
| Great Britain | | | | | | | | | |
| 5 (2005) | H | 556 | yes | yes | 29% | 50% | 27% | 13% | 5% |
| Greece | | | | | | | | | |

| Wave (Year) | Income | n | In SRH data? | In LS data? | TUD | CBC | TUM | %Self | %Self (TU) |
|---|---|---|---|---|---|---|---|---|---|
| 7 (2017) | H | 550 | yes | yes | 15% | 13% | 9% | 15% | 17% |
| Hungary | | | | | | | | | |
| 3 (1998) | UM | 318 | yes | yes | 24% | 38% | 19% | 9% | 2% |
| 5 (2009) | H | 507 | yes | yes | 12% | 25% | 8% | 11% | 2% |
| Indonesia | | | | | | | | | |
| 5 (2006) | LM | 1194 | yes | yes | 10% | 13% | 11% | 28% | 19% |
| 7 (2018) | LM | 2388 | yes | yes | 13% | 10% | 19% | 46% | 33% |
| Italy | | | | | | | | | |
| 5 (2005) | H | 545 | yes | yes | 32% | 100% | 17% | 32% | 18% |
| Japan | | | | | | | | | |
| 1 (1981) | H | 752 | yes | yes | 31% | 27% | 16% | 19% | 2% |
| 2 (1990) | H | 671 | yes | yes | 25% | 22% | 11% | 16% | 3% |
| 3 (1995) | H | 736 | yes | yes | 24% | 21% | 18% | 21% | 2% |
| 4 (2000) | H | 825 | yes | yes | 22% | 19% | 10% | 18% | 6% |
| 5 (2005) | H | 690 | yes | yes | 19% | 16% | 14% | 18% | 9% |
| 7 (2019) | H | 824 | yes | yes | 17% | 15% | 15% | 18% | 6% |
| Kazakhstan | | | | | | | | | |
| 7 (2018) | UM | 831 | yes | yes | 47% | 75% | 20% | 11% | 6% |
| Latvia | | | | | | | | | |
| 3 (1996) | LM | 705 | yes | yes | 29% | 18% | 25% | 8% | 2% |
| Lithuania | | | | | | | | | |
| 3 (1997) | LM | 490 | yes | yes | 19% | 15% | 13% | 13% | 2% |
| Malaysia | | | | | | | | | |
| 5 (2006) | UM | 708 | yes | yes | 11% | 2% | 19% | 20% | 20% |
| 7 (2018) | UM | 973 | yes | yes | 9% | 0% | 37% | 15% | 8% |
| Mexico | | | | | | | | | |
| 2 (1990) | UM | 1011 | yes | yes | 23% | 10% | 5% | 20% | 13% |
| 3 (1996) | UM | 871 | yes | yes | 16% | 12% | 30% | 24% | 24% |
| 4 (2000) | UM | 874 | yes | yes | 16% | 13% | 10% | 26% | 11% |
| 5 (2005) | UM | 838 | yes | yes | 17% | 11% | 20% | 33% | 16% |
| 7 (2018) | UM | 966 | yes | yes | 12% | 10% | 17% | 27% | 22% |
| Moldova | | | | | | | | | |
| 3 (1996) | L | 527 | yes | yes | 28% | 42% | 54% | 5% | 2% |
| 4 (2002) | L | 465 | yes | yes | 28% | 42% | 27% | 11% | 4% |
| 5 (2006) | LM | 532 | yes | yes | 28% | 42% | 28% | 16% | 9% |
| Montenegro | | | | | | | | | |
| 3 (1996) | LM | 129 | yes | yes | 26% | 75% | 43% | 13% | 5% |
| 4 (2001) | LM | 495 | yes | yes | 26% | 75% | 30% | 16% | 8% |
| Morocco | | | | | | | | | |
| 4 (2001) | LM | 911 | yes | yes | 6% | 3% | 1% | 50% | 0% |
| 5 (2007) | LM | 1040 | yes | yes | 6% | 3% | 9% | 30% | 12% |
| Netherlands | | | | | | | | | |
| 5 (2006) | H | 518 | yes | yes | 20% | 67% | 25% | 6% | 4% |
| 6 (2012) | H | 924 | yes | yes | 18% | 81% | 22% | 5% | 1% |
| New Zealand | | | | | | | | | |
| 3 (1998) | H | 713 | yes | yes | 22% | 30% | 22% | 0% | 0% |
| 5 (2004) | H | 597 | yes | yes | 22% | 20% | 23% | 0% | 0% |

| Wave (Year) | Income | n | In SRH data? | In LS data? | TUD | CBC | TUM | %Self | %Self (TU) |
|---|---|---|---|---|---|---|---|---|---|
| North Macedonia | | | | | | | | | |
| 3 (1998) | LM | 471 | yes | yes | 28% | 49% | 28% | 13% | 11% |
| 4 (2001) | LM | 429 | yes | yes | 28% | 49% | 11% | 9% | 6% |
| Norway | | | | | | | | | |
| 3 (1996) | H | 819 | yes | yes | 56% | 77% | 57% | 13% | 4% |
| 5 (2007) | H | 717 | yes | yes | 50% | 74% | 56% | 10% | 6% |
| Peru | | | | | | | | | |
| 3 (1996) | LM | 569 | yes | yes | 2% | 5% | 20% | 36% | 31% |
| 4 (2001) | LM | 731 | yes | yes | 2% | 5% | 5% | 32% | 23% |
| 7 (2018) | UM | 909 | yes | yes | 5% | 5% | 5% | 45% | 26% |
| Philippines | | | | | | | | | |
| 3 (1996) | LM | 630 | yes | yes | 10% | 4% | 12% | 38% | 29% |
| 4 (2001) | LM | 608 | yes | yes | 10% | 4% | 5% | 39% | 18% |
| 7 (2019) | LM | 712 | yes | yes | 8% | 1% | 22% | 43% | 40% |
| Poland | | | | | | | | | |
| 5 (2005) | UM | 441 | yes | yes | 25% | 19% | 20% | 8% | 2% |
| Romania | | | | | | | | | |
| 3 (1998) | LM | 680 | yes | yes | 46% | 100% | 31% | 12% | 3% |
| 5 (2005) | UM | 719 | yes | yes | 34% | 100% | 15% | 6% | 2% |
| 7 (2018) | UM | 537 | yes | yes | 21% | 48% | 17% | 8% | 7% |
| Russian Federation | | | | | | | | | |
| 2 (1990) | UM | 1350 | yes | yes | 43% | 28% | 76% | 0% | 0% |
| 3 (1995) | LM | 1185 | yes | yes | 43% | 28% | 58% | 2% | 0% |
| 5 (2006) | UM | 1194 | yes | yes | 43% | 28% | 23% | 1% | 0% |
| 7 (2017) | UM | 1080 | yes | yes | 27% | 30% | 10% | 3% | 2% |
| Rwanda | | | | | | | | | |
| 5 (2007) | L | 885 | yes | yes | 5% | 5% | 28% | 78% | 69% |
| Serbia | | | | | | | | | |
| 3 (1996) | LM | 648 | yes | yes | 31% | 55% | 44% | 10% | 2% |
| 4 (2001) | LM | 616 | yes | yes | 31% | 55% | 25% | 12% | 4% |
| 5 (2006) | UM | 643 | yes | yes | 28% | 55% | 23% | 10% | 6% |
| 7 (2017) | UM | 505 | yes | yes | 22% | 30% | 20% | 9% | 3% |
| Singapore | | | | | | | | | |
| 4 (2002) | H | 864 | NO | yes | 19% | 18% | 8% | 4% | 0% |
| Slovakia | | | | | | | | | |
| 3 (1998) | UM | 635 | yes | yes | 40% | 52% | 31% | 3% | 1% |
| Slovenia | | | | | | | | | |
| 3 (1995) | UM | 536 | yes | yes | 50% | 100% | 50% | 6% | 3% |
| 5 (2005) | H | 505 | yes | yes | 37% | 100% | 35% | 6% | 1% |
| South Africa | | | | | | | | | |
| 3 (1996) | UM | 1439 | yes | yes | 30% | 46% | 35% | 9% | 6% |
| 4 (2001) | LM | 1315 | yes | yes | 32% | 46% | 12% | 11% | 4% |
| 5 (2006) | UM | 1282 | yes | yes | 30% | 42% | 35% | 12% | 7% |
| South Korea | | | | | | | | | |
| 3 (1996) | H | 802 | yes | NO | 12% | 15% | 12% | 44% | 41% |
| 4 (2001) | H | 726 | yes | yes | 12% | 15% | 8% | 30% | 18% |
| 7 (2018) | H | 857 | yes | yes | 12% | 15% | 8% | 12% | 11% |

| Wave (Year) | Income | n | In SRH data? | In LS data? | TUD | CBC | TUM | %Self | %Self (TU) |
|---|---|---|---|---|---|---|---|---|---|
| Spain | | | | | | | | | |
| 2 (1990) | H | 602 | yes | yes | 13% | 83% | 8% | 11% | 10% |
| 3 (1995) | H | 430 | yes | yes | 16% | 83% | 19% | 10% | 5% |
| 4 (2000) | H | 552 | yes | yes | 15% | 83% | 7% | 13% | 3% |
| Sweden | | | | | | | | | |
| 3 (1996) | H | 663 | yes | yes | 83% | 90% | 76% | 7% | 2% |
| 4 (1999) | H | 661 | NO | yes | 81% | 88% | 72% | 3% | 2% |
| 5 (2006) | H | 669 | yes | yes | 74% | 89% | 69% | 7% | 3% |
| Switzerland | | | | | | | | | |
| 2 (1989) | H | 819 | yes | yes | 23% | 49% | 9% | 8% | 0% |
| 3 (1996) | H | 757 | yes | yes | 23% | 46% | 20% | 13% | 9% |
| Thailand | | | | | | | | | |
| 5 (2007) | LM | 1194 | yes | yes | 3% | 1% | 17% | 64% | 56% |
| 7 (2018) | UM | 1203 | yes | yes | 3% | 2% | 35% | 60% | 67% |
| Trinidad and Tobago | | | | | | | | | |
| 5 (2006) | H | 524 | yes | yes | 21% | 20% | 24% | 21% | 12% |
| Tunisia | | | | | | | | | |
| 7 (2019) | LM | 574 | yes | yes | 38% | 63% | 9% | 34% | 32% |
| Turkey | | | | | | | | | |
| 2 (1990) | LM | 422 | yes | yes | 25% | 25% | 0% | 35% | - |
| 3 (1996) | LM | 848 | yes | yes | 16% | 16% | 9% | 32% | 8% |
| 5 (2007) | UM | 526 | yes | yes | 9% | 8% | 5% | 34% | 15% |
| 6 (2011) | UM | 665 | yes | yes | 8% | 7% | 4% | 20% | 4% |
| 7 (2018) | UM | 1349 | yes | yes | 9% | 10% | 8% | 17% | 11% |
| Uganda | | | | | | | | | |
| 4 (2001) | L | 638 | yes | yes | 11% | 15% | 10% | 36% | 32% |
| Ukraine | | | | | | | | | |
| 3 (1996) | LM | 1553 | yes | yes | 57% | 55% | 47% | 5% | 2% |
| 5 (2006) | LM | 555 | yes | yes | 57% | 55% | 30% | 7% | 4% |
| United States | | | | | | | | | |
| 3 (1995) | H | 889 | yes | yes | 15% | 17% | 26% | 8% | 4% |
| 4 (1999) | H | 831 | yes | yes | 14% | 15% | 14% | 7% | 2% |
| 5 (2006) | H | 710 | yes | yes | 12% | 13% | 19% | 16% | 4% |
| 7 (2017) | H | 1730 | yes | yes | 11% | 12% | 19% | 8% | 5% |
| Uruguay | | | | | | | | | |
| 3 (1996) | UM | 493 | yes | yes | 17% | 89% | 17% | 18% | 13% |
| 5 (2006) | UM | 496 | yes | yes | 17% | 89% | 10% | 25% | 6% |
| Venezuela | | | | | | | | | |
| 3 (1996) | LM | 645 | yes | yes | 0% | 8% | 19% | 33% | 17% |
| 4 (2000) | UM | 630 | NO | yes | 0% | 8% | 4% | 32% | 4% |
| Vietnam | | | | | | | | | |
| 4 (2001) | L | 329 | yes | yes | 14% | 25% | 20% | 37% | 9% |
| 5 (2006) | L | 664 | yes | yes | 14% | 25% | 16% | 43% | 18% |
| Zambia | | | | | | | | | |
| 5 (2007) | L | 487 | yes | yes | 44% | 38% | 38% | 30% | 18% |

### Availability of TUD/CBC

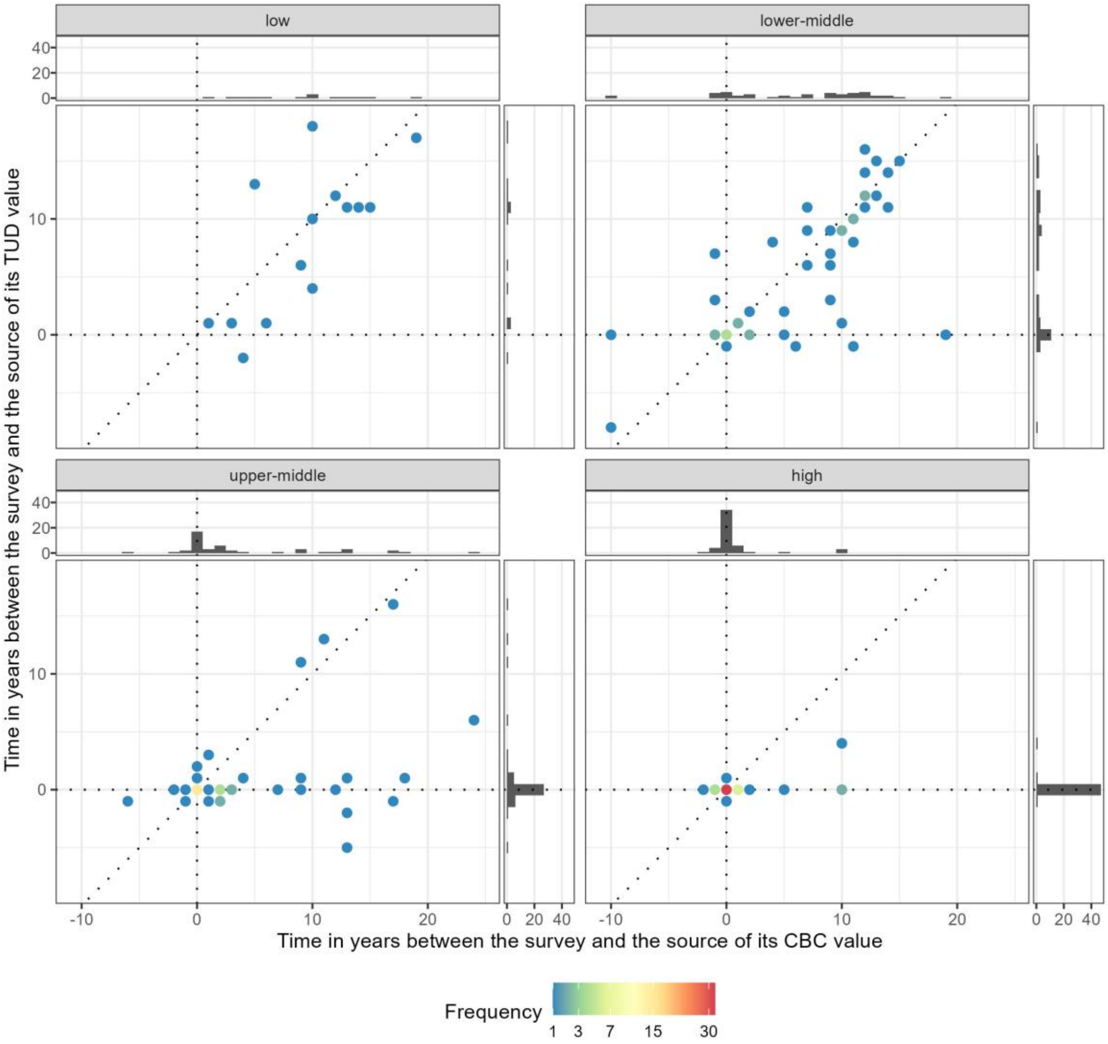


Figure S2: Time between survey and source of corresponding TUD/CBC values by country income. Horizontal/vertical dotted lines indicate 0, i.e., that the value was from the same year. Diagonal dotted lines indicate x = y. Negative values indicate that TUD/CBC values from the past were used, while positive values represent values from the future. Histograms in the margins show distribution of individual variables.

### Table with values in Figure 2

Table S2: Values plotted in Figure 2

| | Self-rated health | Life satisfaction |
|---|---|---|
| TUD (1985) | | |
| low/lower-middle only | 1.18 (0.94-1.47) | 1.13 (0.90-1.42) |
| + country-lvl | 1.17 (1.10-1.25) | 1.02 (0.96-1.09) |
| + expo-related | 1.19 (1.12-1.27) | 1.03 (0.97-1.10) |
| + indv-lvl | 1.31 (1.24-1.38) | 1.17 (1.11-1.24) |
| expo*year | 1.33 (1.26-1.40) | 1.17 (1.11-1.23) |
| TUD (1995) | | |
| low/lower-middle only | 0.93 (0.75-1.14) | 0.87 (0.71-1.08) |
| + country-lvl | 1.05 (0.98-1.12) | 0.93 (0.87-0.99) |
| + expo-related | 1.10 (1.04-1.17) | 0.96 (0.90-1.02) |
| + indv-lvl | 1.22 (1.17-1.28) | 1.10 (1.05-1.16) |
| expo*year | 1.21 (1.16-1.27) | 1.09 (1.04-1.14) |
| TUD (2005) | | |
| low/lower-middle only | 0.73 (0.59-0.90) | 0.67 (0.55-0.84) |
| + country-lvl | 0.94 (0.87-1.00) | 0.84 (0.79-0.90) |
| + expo-related | 1.02 (0.95-1.09) | 0.89 (0.83-0.95) |
| + indv-lvl | 1.14 (1.09-1.20) | 1.04 (0.99-1.09) |
| expo*year | 1.11 (1.06-1.17) | 1.01 (0.96-1.06) |
| TUD (2015) | | |
| low/lower-middle only | 0.57 (0.46-0.72) | 0.52 (0.41-0.66) |
| + country-lvl | 0.84 (0.77-0.90) | 0.77 (0.71-0.83) |
| + expo-related | 0.94 (0.87-1.01) | 0.82 (0.76-0.89) |
| + indv-lvl | 1.07 (1.01-1.13) | 0.98 (0.93-1.04) |
| expo*year | 1.02 (0.97-1.07) | 0.94 (0.89-0.99) |
| CBC (1985) | | |
| low/lower-middle only | 2.70 (1.58-4.61) | 3.06 (1.80-5.19) |
| + country-lvl | 1.29 (1.18-1.41) | 1.27 (1.16-1.38) |
| + expo-related | 1.12 (1.03-1.21) | 1.18 (1.09-1.28) |
| + indv-lvl | 1.24 (1.17-1.32) | 1.17 (1.10-1.24) |
| expo*year | 1.21 (1.14-1.28) | 1.16 (1.09-1.23) |
| CBC (1995) | | |
| low/lower-middle only | 1.84 (1.10-3.08) | 2.08 (1.25-3.46) |
| + country-lvl | 1.26 (1.16-1.36) | 1.26 (1.17-1.37) |
| + expo-related | 1.11 (1.03-1.20) | 1.19 (1.11-1.28) |
| + indv-lvl | 1.22 (1.16-1.29) | 1.19 (1.12-1.25) |
| expo*year | 1.19 (1.12-1.25) | 1.16 (1.10-1.22) |
| CBC (2005) | | |
| low/lower-middle only | 1.25 (0.76-2.08) | 1.42 (0.86-2.33) |
| + country-lvl | 1.22 (1.14-1.32) | 1.26 (1.17-1.36) |
| + expo-related | 1.11 (1.04-1.20) | 1.20 (1.12-1.29) |
| + indv-lvl | 1.20 (1.14-1.27) | 1.20 (1.13-1.27) |
| expo*year | 1.16 (1.10-1.23) | 1.16 (1.10-1.22) |
| CBC (2015) | | |
| low/lower-middle only | 0.86 (0.51-1.43) | 0.96 (0.58-1.60) |

| | Self-rated health | Life satisfaction |
|---|---|---|
| + country-lvl | 1.19 (1.11-1.29) | 1.26 (1.17-1.36) |
| + expo-related | 1.11 (1.03-1.19) | 1.22 (1.13-1.31) |
| + indv-lvl | 1.19 (1.11-1.26) | 1.21 (1.14-1.29) |
| expo*year | 1.14 (1.07-1.21) | 1.16 (1.09-1.23) |

### Predicted expected outcome in lower-middle income countries

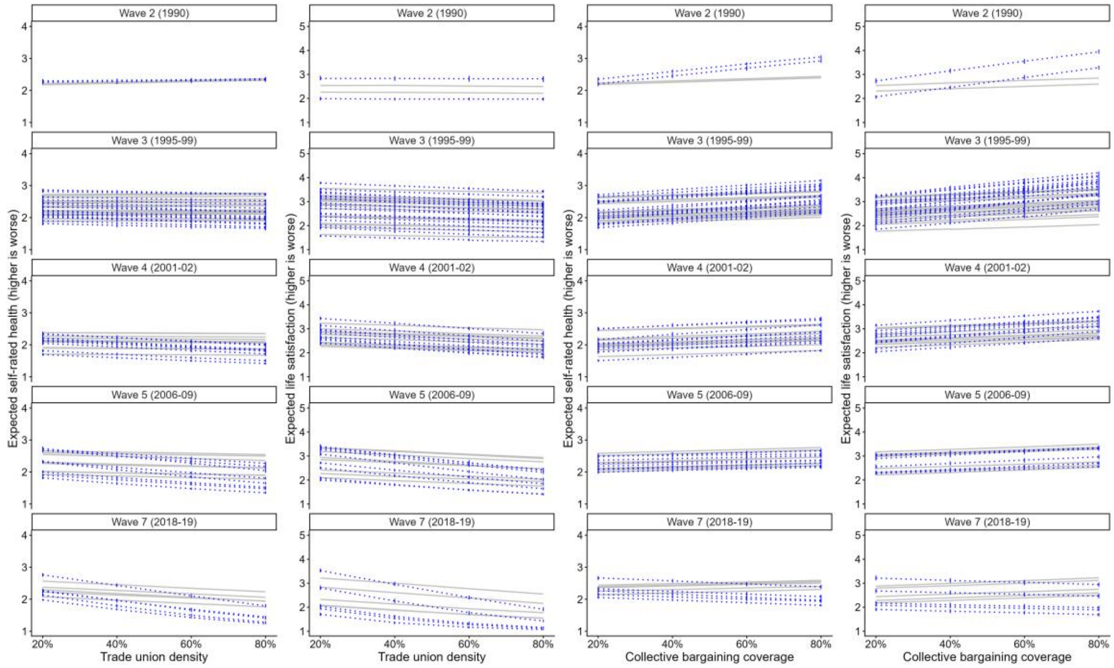


Figure S3: Predicted expected self-rated health (columns 1 & 3) and life satisfaction (columns 2 & 4) for different values of trade union density (column 1 & 2) and collective bargaining coverage (columns 3 & 4) by country (lower-middle income only) and study wave in fully adjusted models including all countries (blue dotted lines) and including low and lower-middle income countries only (grey lines). Line ranges show 2.5 and 97.5% quantiles for predicted expected outcomes, for exposure values of 20%, 40%, 60%, and 80%. Lines only connect these four sets of predictions.

### Sensitivity analyses

#### Including surveys after 2019

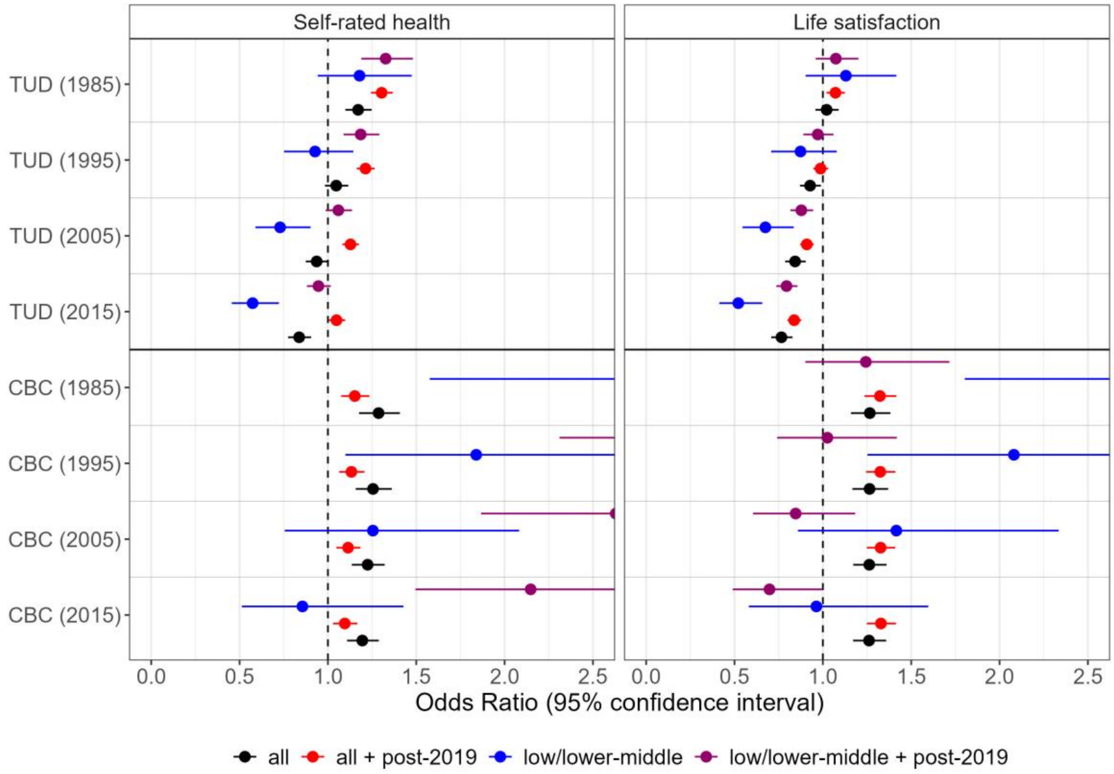


Figure S4: Comparison of effect estimates from fully adjusted models in all countries (all) and in low and lower-middle income countries only (low/lower-middle) with the corresponding models that do not exclude surveys after 2019 (+ post-2019)

Restrict to surveys with TUD/CBC available for the same year

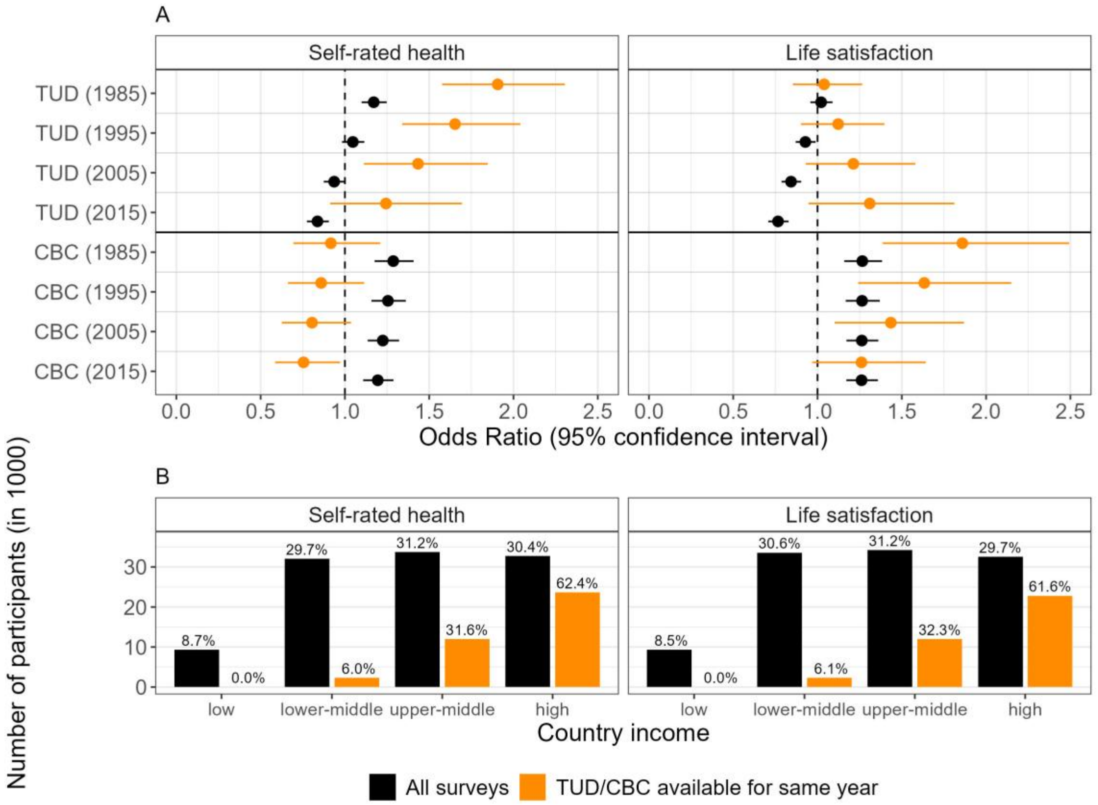


Figure S5: Results from models including only surveys for which TUD/CBC are available for the survey year. (A) Comparison of effect estimates of fully adjusted models in all surveys (All surveys) and fully adjusted restricted models (TUD/CBC available for same year). (B) Number of participants included in the models by country income. Percentages describe proportions of country income category among all participants.

Restrict to employees

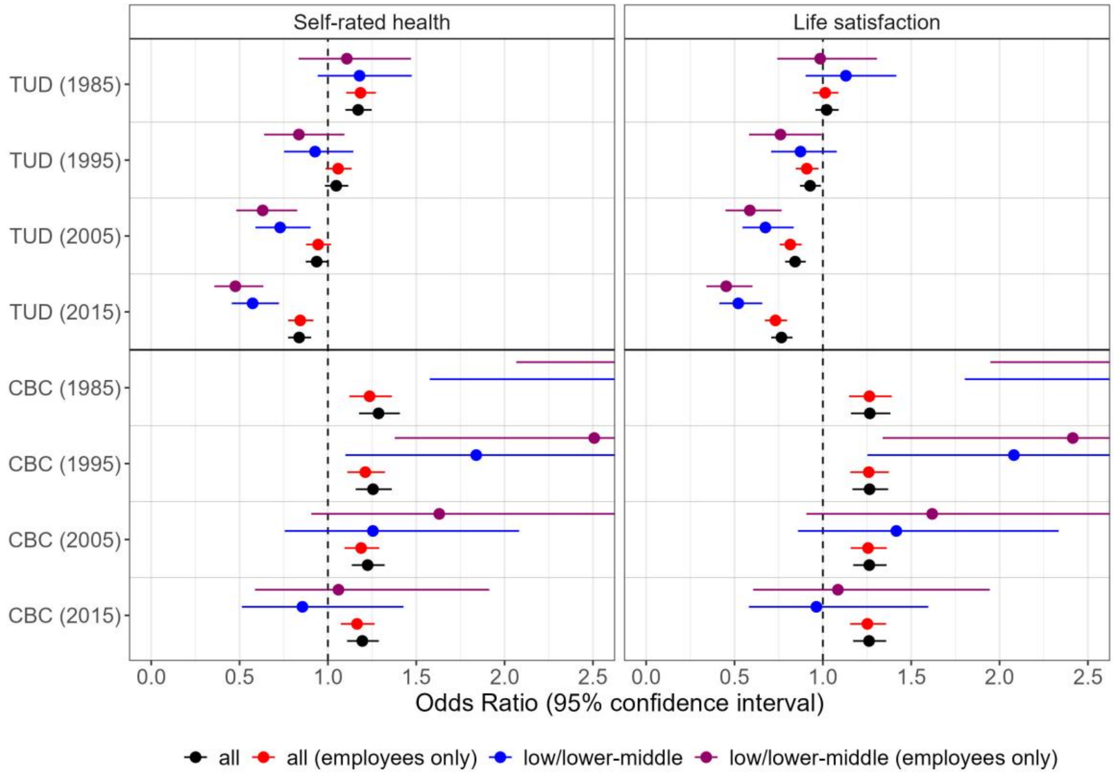


Figure S6: Comparison of effect estimates from fully adjusted models in all countries (all) and in low and lower-middle income countries only (low/lower-middle) with the corresponding models that only include employed (full-time and part-time) and exclude self-employed (employees only)

Weighted analysis

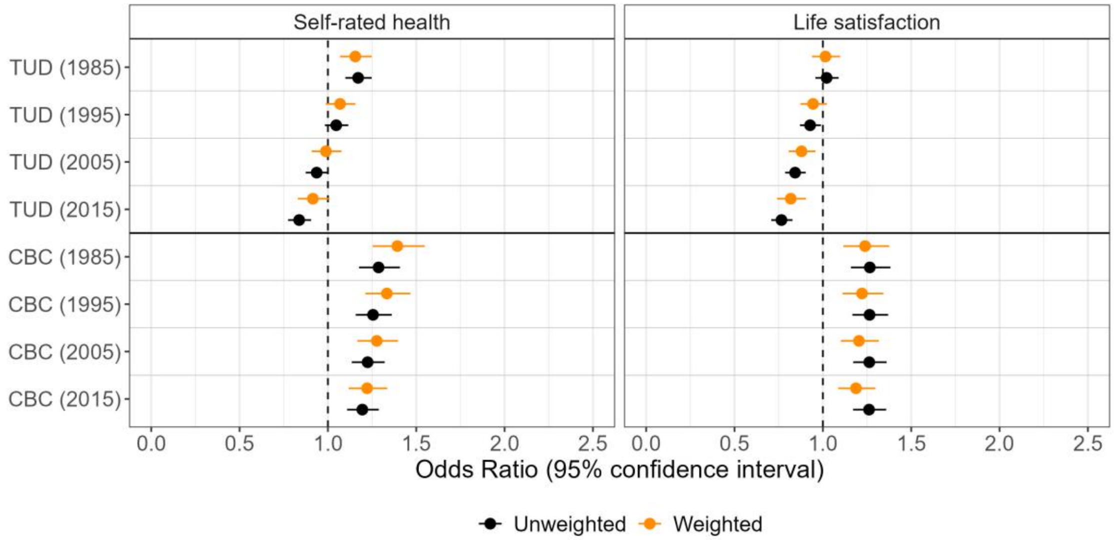


Figure S7: Comparison of effect estimates from fully adjusted unweighted models in all countries (Unweighted) and the corresponding weighted models (Weighted) that weigh all countries equally

### Stratified analysis versus interaction effects

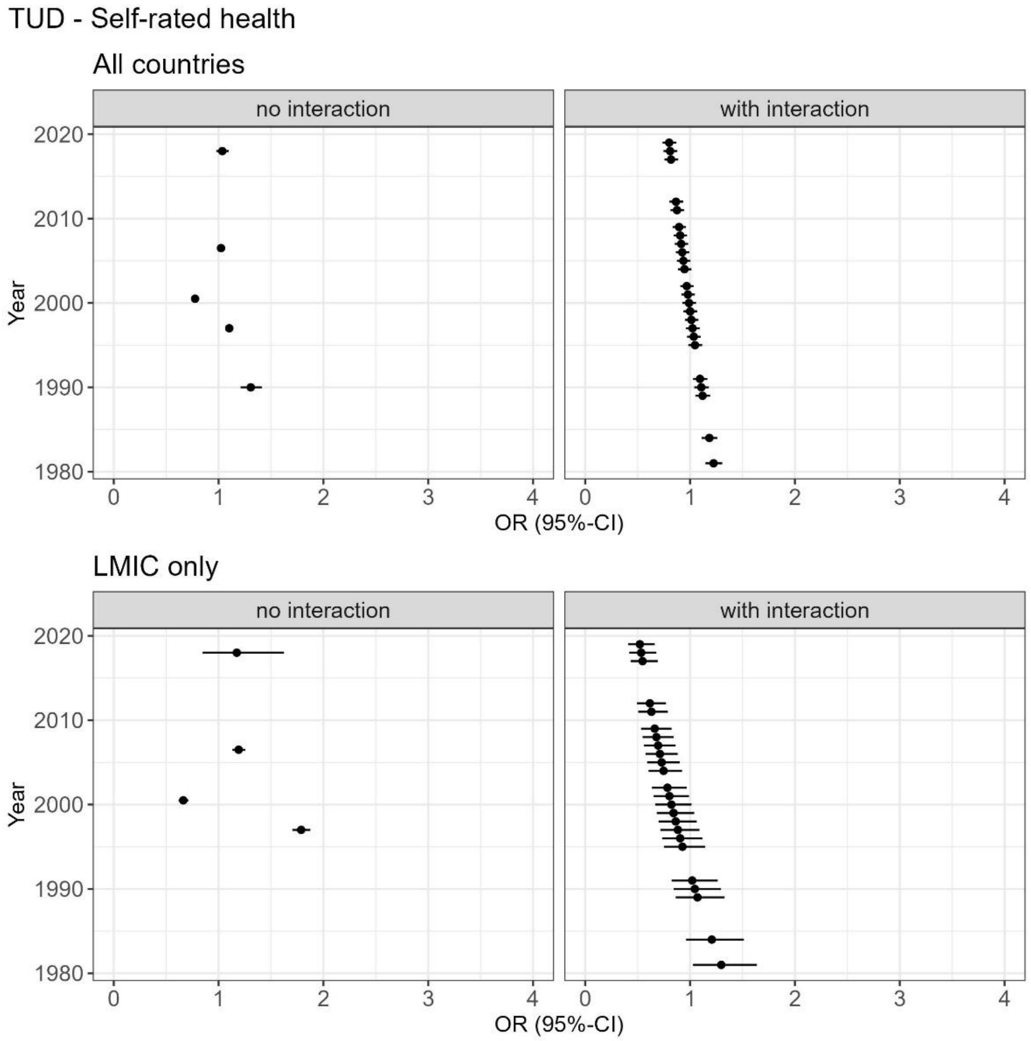


Figure S8a: Comparison of effect estimates for trade union density (TUD) on self-rated health from fully adjusted models without interaction with survey year but stratified by wave (no interaction), and the main models (with interaction). Comparisons are shown for models including data from all countries ("All countries", top row) and models including only data from low- and lower-middle income countries ("LMIC only", bottom row). In "no interaction" panels, estimates represent a complete wave with data from multiple years, but they are plotted at the average year for each wave. Some waves are missing due to low amount of data. In "with interaction" panels, estimates from the main model have been calculated for all years in which included surveys have been conducted (while previous plots only show estimates for years 1985, 1995, 2005, and 2015).

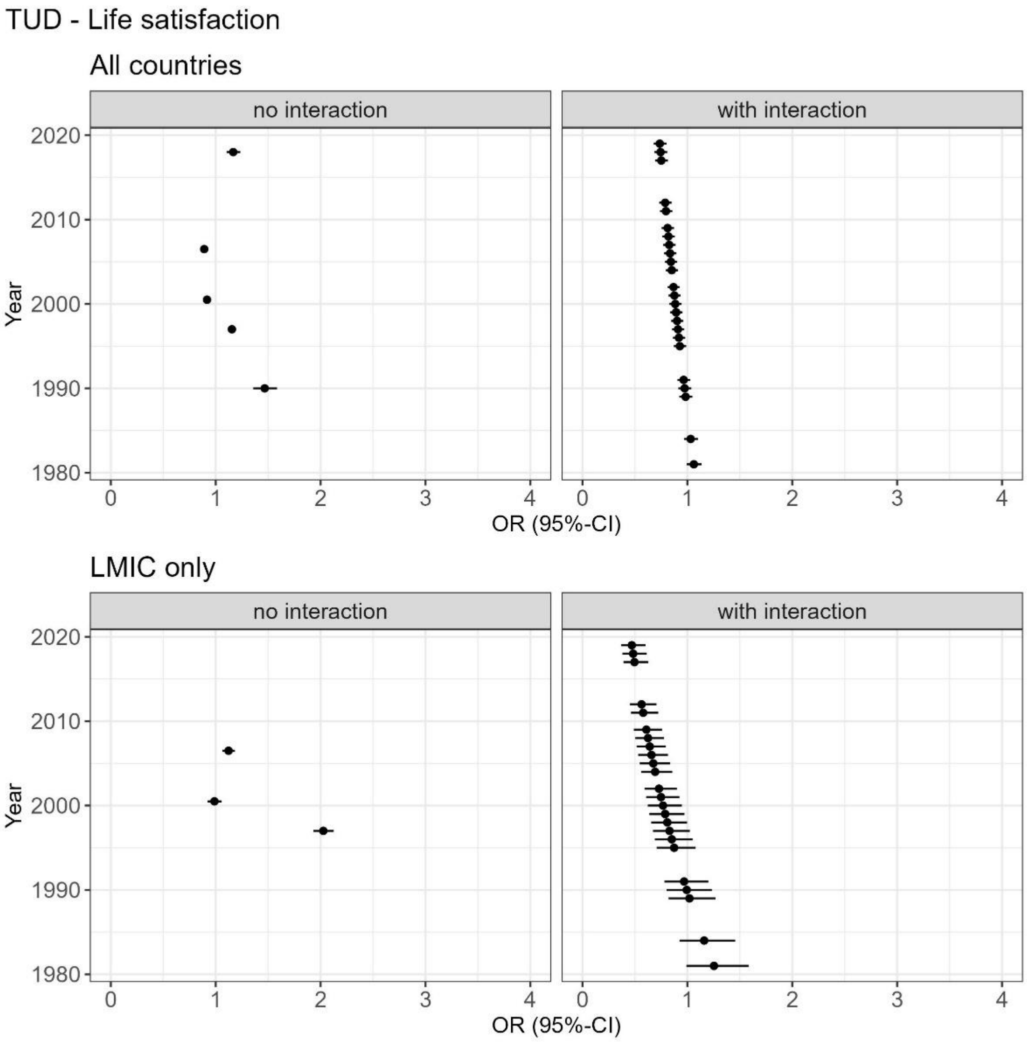


Figure S8b: Comparison of effect estimates for trade union density (TUD) on life satisfaction from fully adjusted models without interaction with survey year but stratified by wave (no interaction), and the main models (with interaction). Comparisons are shown for models including data from all countries ("All countries", top row) and models including only data from low- and lower-middle income countries ("LMIC only", bottom row). In "no interaction" panels, estimates represent a complete wave with data from multiple years, but they are plotted at the average year for each wave. Some waves are missing due to low amount of data. In "with interaction" panels, estimates from the main model have been calculated for all years in which included surveys have been conducted (while previous plots only show estimates for years 1985, 1995, 2005, and 2015).

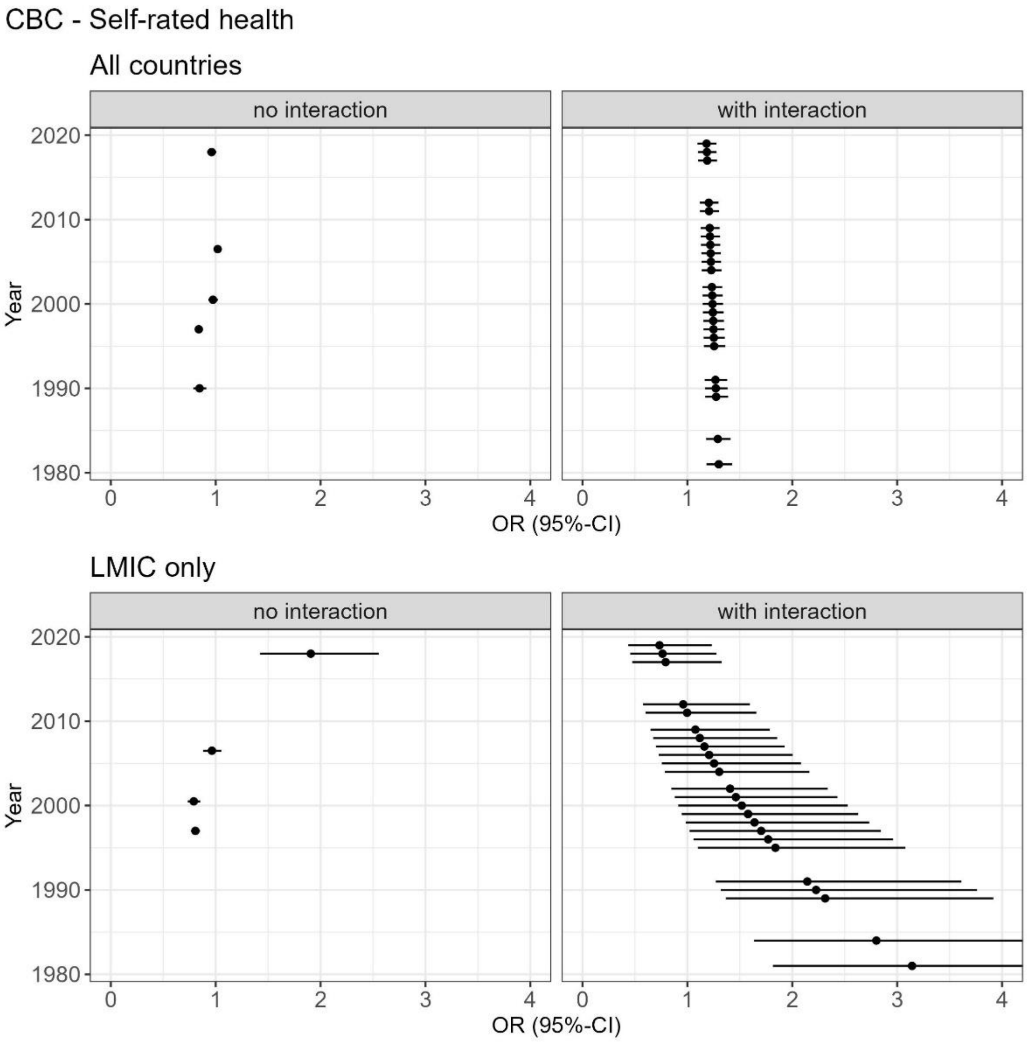


Figure S8c: Comparison of effect estimates for collective bargaining coverage (CBC) on self-rated health from fully adjusted models without interaction with survey year but stratified by wave (no interaction), and the main models (with interaction). Comparisons are shown for models including data from all countries ("All countries", top row) and models including only data from low- and lower-middle income countries ("LMIC only", bottom row). In "no interaction" panels, estimates represent a complete wave with data from multiple years, but they are plotted at the average year for each wave. Some waves are missing due to low amount of data. In "with interaction" panels, estimates from the main model have been calculated for all years in which included surveys have been conducted (while previous plots only show estimates for years 1985, 1995, 2005, and 2015).

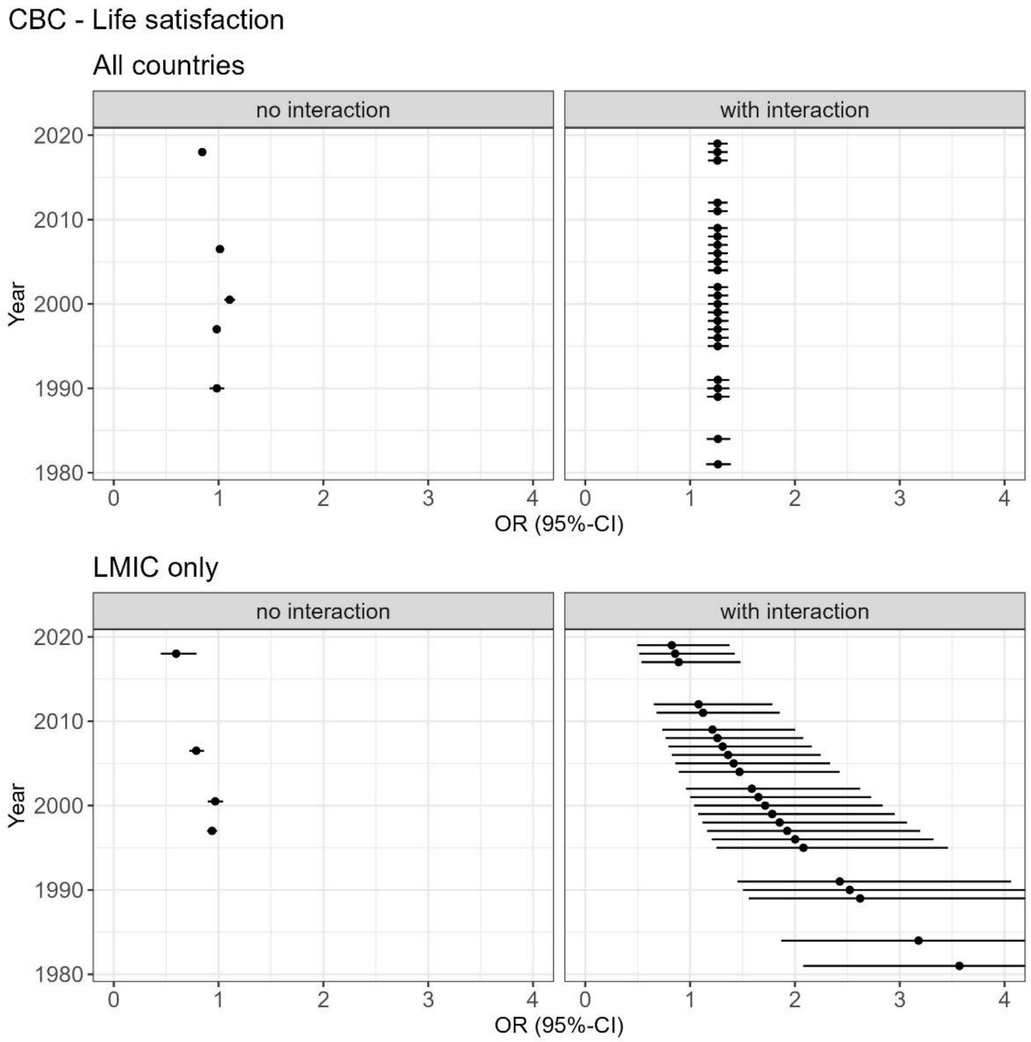


Figure S8d: Comparison of effect estimates for collective bargaining coverage (CBC) on life satisfaction from fully adjusted models without interaction with survey year but stratified by wave (no interaction), and the main models (with interaction). Comparisons are shown for models including data from all countries ("All countries”, top row) and models including only data from low- and lower-middle income countries (“LMIC only”, bottom row). In “no interaction” panels, estimates represent a complete wave with data from multiple years, but they are plotted at the average year for each wave. Some waves are missing due to low amount of data. In “with interaction” panels, estimates from the main model have been calculated for all years in which included surveys have been conducted (while previous plots only show estimates for years 1985, 1995, 2005, and 2015).
